# Beyond event-rate enrichment: proteomic risk scores for mechanism-aware prevention trial design

**DOI:** 10.64898/2026.06.09.26355266

**Authors:** Joshua Fieggen, Gregory Simond, Bradley Max Segal, Ayush Noori, Anjan Thakurta, Christopher C. Butler, David A. Clifton, Lei Clifton

## Abstract

**Background:** Blood-based biomarkers are increasingly proposed for identifying high-risk individuals before clinical disease and for making prevention-oriented trials more efficient. Prognostic enrichment can increase event rates, but trial efficiency also depends on whether the intervention effect is preserved in the enriched population.

**Methods:** Using the UK Biobank Pharma Proteomics Project, we trained disease-specific proteomic risk scores (ProRS) from 2,916 plasma proteins with elastic-net Cox models. We compared ProRS, polygenic risk scores (PRS), and combined PRS–ProRS scores across ten incident diseases. We estimated cumulative incidence and theoretical two-arm time-to-event trial sample sizes across risk strata. To evaluate effect preservation, we examined six intervention-analogue exposure–outcome pairs spanning genetic (*PCSK9* /coronary artery disease, *APOE* /Alzheimer’s disease, *PPARG*/type 2 diabetes, *IL23R*/Crohn’s disease), behavioural (physical activity/all-cause mortality), and pharmacological (RAAS inhibitors versus calcium channel blockers/coronary artery disease) examples.

**Results:** ProRS outperformed PRS for 9 of 10 diseases (median C-index 0.75 versus 0.61). ProRS and PRS were weakly correlated (median Pearson |*r*| = 0.04), and joint PRS–ProRS stratification identified groups with higher observed incidence than either score alone for several endpoints. In the top risk quartile, combined-score enrichment reduced theoretical required sample sizes by 32–74% under a fixed 20% relative hazard reduction. These gains were not always preserved when stratum-specific intervention-analogue effects were used. Effects were broadly preserved for *APOE* /Alzheimer’s disease and physical activity/mortality. The *PPARG*/type 2 diabetes effect attenuated toward the null under all three score types, showing that event-rate enrichment does not guarantee effect preservation. For *IL23R*/Crohn’s disease and the antihypertensive comparison, point estimates differed across score types – preserved under polygenic but attenuated under proteomic enrichment – but confidence intervals were wide and overlapping.

**Conclusions:** Proteomic risk scores can identify high-event-rate populations for prevention-oriented trials, but event-rate enrichment alone is insufficient for trial design. Biomarker-guided enrichment should evaluate mechanism-specific effect preservation and may be preferable as a stratification or adaptive-design variable rather than as a restrictive eligibility criterion.

## 1 Introduction

Prevention-oriented trials often require large populations and long follow-up because clinically meaningful endpoints are uncommon in unselected participants [1, 2]. Prognostic enrichment addresses this problem by preferentially enrolling individuals at higher baseline risk, thereby increasing the expected number of events for a given trial size [3–6]. Regulatory guidance recognises enrichment strategies, including biomarker- and risk-score-based selection, as a way to improve the efficiency of trials when the target population is well defined [7].

The feasibility of such designs is changing because scalable blood-based biomarkers can identify biological risk before clinical disease. Plasma proteomic platforms now measure thousands of circulating proteins at scale [8], and proteomic risk scores have shown strong prediction for many common and rare diseases [9– 12]. Disease-specific plasma biomarkers are also increasingly discussed as tools for early detection and trial targeting in preclinical disease states; for example, plasma Alzheimer’s disease neuropathology biomarkers can now be detected in midlife and are associated with worse cognition and accelerated decline well before clinical disease [13]. These developments raise the question of whether proteomic biomarkers can be used to identify populations in which prevention trials are smaller, faster, or more informative.

Polygenic risk scores (PRS) provide one model for this approach. PRS quantify inherited susceptibility, can be measured once, and have been proposed as trial-enrichment tools [14–17]. Cai *et al*. recently described an in silico framework for PRS-guided prognostic enrichment, using naturally occurring protective genetic variants as therapeutic analogues and showing that PRS restriction can increase disease prevalence, improve power, reduce sample size, and accelerate event accrual [18]. However, much of the growing interest is focused on phenotypic biomarkers such as proteomics, as these have been shown to outperform genetic biomarkers in risk prediction [TODO cite milton].

In addition, theoretical efficiency gains assume that the relative treatment effect is constant across baseline-risk strata [5, 7]. This assumption has clear limits – in COVID-19, for example, remdesivir appeared more effective in lower-risk than higher-risk patients [19]. A phenotypic biomarker is more likely to identify people with subclinical disease, downstream organ damage, inflammation, or metabolic change. Such individuals may have higher event rates but may not respond similarly to an intervention that acts earlier in the disease pathway. Event-rate enrichment could therefore overstate trial efficiency if the intervention effect is attenuated in the selected population.

We evaluated this trade-off in the UK Biobank. We trained proteomic risk scores (ProRS) for ten incident diseases, compared them with PRS and combined PRS–ProRS scores, estimated theoretical sample-size reductions under prognostic enrichment, and assessed whether intervention-analogue effects were preserved across enrichment strata.

## 2 Methods

### 2.1 Study population and proteomics data

The UK Biobank (UKB) is a prospective cohort of approximately 500,000 participants aged 40–69 years recruited between 2006 and 2010 [20]. Baseline data were collected through questionnaires, interviews, physical measures, and biological samples, with follow-up through linked electronic health records, death records, cancer registries, and hospital inpatient records. All participants provided written informed consent and UKB has ethical approval from the National Health Service Northwest Multicentre Research Ethics Committee (06/MRE08/65).

The UK Biobank Pharma Proteomics Project (UKB-PPP) measured 2,923 plasma proteins in more than 54,000 participants using the Olink Proximity Extension Assay [8]. After UKB-PPP quality control, we excluded non-baseline samples, samples missing more than 1,000 proteins, and seven proteins with more than 10% missingness, leaving 2,916 proteins in 44,802 participants. Participants were randomly split 50:50 into training and test sets. Proteins were standardised using training-set means and standard deviations; missing values were imputed with *k*-nearest neighbours using 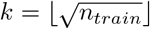; and test data were transformed using parameters learned in the training data. Internal masking of 3% of observed values gave a mean absolute imputation error of 0.3280 in training data and 0.3279 in test data.

### 2.2 Disease endpoints

We defined ten incident endpoints: coronary artery disease (CAD), type 2 diabetes (T2D), atrial fibrillation (AF), breast cancer (BC; females only), colorectal cancer (CRC), prostate cancer (PC; males only), Crohn’s disease (CD), venous thromboembolism (VTE), ischaemic stroke (ISS), and Alzheimer’s disease (AD). End-point definitions used ICD-10 and ICD-9 codes from hospital records, cancer registry data, and death records (Supplementary Table S1). Participants with linked-record or self-reported disease on or before 90 days after baseline were excluded as prevalent cases. Follow-up ran from baseline until first diagnosis, death, loss to follow-up, or administrative censoring on 31 May 2022.

### 2.3 Risk score construction and evaluation

Disease-specific PRS were obtained from UK Biobank Category 300, using the externally trained Standard PRS release described by Thompson *et al*. [16]. PRS values were *z*-standardised within the study population.

For each disease, we trained an elastic-net Cox proportional hazards model on the 2,916 proteins in the training set, using no clinical covariates. The elastic-net mixing parameter was fixed at 0.5 and the regularisation strength was chosen by five-fold stratified cross-validation to maximise Harrell’s C-index. ProRS were calculated as the model linear predictor and *z*-standardised using the training-set distribution. Combined scores were constructed by fitting an unregularised Cox model with PRS and ProRS as predictors in the training set, then applying the fitted linear combination to all participants. Where PRS was missing but ProRS was available (*∼*1%), PRS was set to zero, corresponding to the population mean after standardisation. Disease-specific ProRS coefficients estimated in the training set are listed in Supplementary Table S12

Predictive discrimination was assessed in the held-out test set using Harrell’s C-index. Observed 10-year cumulative incidence was also examined across score deciles and quartiles using Kaplan-Meier estimates.

### 2.4 Enrichment and theoretical sample size

For each disease and score type, we defined enriched strata corresponding to the top 75%, 50%, 25%, 10%, and 5% of the score distribution, with the full test population as the reference. Within each stratum, we estimated 5- and 10-year cumulative incidence using Kaplan-Meier methods.

We then estimated the number of participants required for a hypothetical two-arm time-to-event trial within each stratum. Calculations used an event-driven log-rank design under proportional hazards and the Schoenfeld formula [21]; full equations are given in Supplementary Methods S1. The primary scenario assumed 1:1 randomisation, two-sided *α* = 0.05, 80% power, a 10-year endpoint, and a target hazard ratio of 0.80. The control-arm event risk was set to the observed Kaplan-Meier cumulative incidence within the stratum, and the treatment-arm risk was derived under proportional hazards. Sample-size reductions were expressed relative to the unselected population. These calculations are theoretical upper bounds: they assume instantaneous enrolment, no additional loss to follow-up, no screening or assay cost, and a constant relative treatment effect unless otherwise stated. Confidence intervals were obtained from 300 bootstraps.

### 2.5 Joint PRS–ProRS stratification

To assess complementarity between inherited and proteomic risk, we calculated Pearson correlations between PRS and ProRS for each disease. We also created four groups using the top-quartile thresholds of both scores: low PRS/low ProRS, low PRS/high ProRS, high PRS/low ProRS, and high PRS/high ProRS. Ten-year cumulative incidence was estimated within each group.

### 2.6 Intervention-analogue effect preservation

We evaluated whether enrichment changed estimated intervention-analogue effects across six exposure– outcome pairs. This extended the protective-variant framework used by Cai *et al*. [18] from PRS-only enrichment to PRS, ProRS, and Combined enrichment, and added behavioural and pharmacological examples.

For genetic therapeutic analogues, carriers of protective variants were compared with non-carriers for four gene–disease pairs: *PCSK9* p.Arg46Leu (rs11591147) for CAD [22], *APOE* p.Arg158Cys (rs7412; proxy for *APOE ε*2) for AD [23], *PPARG* p.Pro12Ala (rs1801282) for T2D [24], and *IL23R* p.Arg381Gln (rs11209026) for CD [25]. Carrier status was derived from imputed genotypes [26]. Cox models estimated the carrier effect within the full population and within the top 75%, 50%, and 25% strata for each score, adjusting for age and sex. Both training and test sets were used because the objective was effect estimation rather than predictive validation.

For a behavioural analogue, we compared high (top tertile) versus low (bottom tertile) accelerometer-measured moderate-to-vigorous physical activity (MVPA) for all-cause mortality within CAD enrichment strata [27]. A two-year washout excluded deaths soon after accelerometer assessment. Cox models adjusted for age at accelerometry, sex, and season of wear. For a pharmacological example, we used a new-user active-comparator design comparing renin–angiotensin–aldosterone system inhibitors with calcium channel blockers for incident CAD. Stabilised inverse-probability weights were estimated from age, sex, self-reported ethnicity, systolic blood pressure, estimated glomerular filtration rate, diabetes status, and body mass index, and truncated at the 1st and 99th percentiles before weighted Cox modelling.

### 2.7 Sensitivity analyses and reproducibility

Sensitivity analyses compared PRS, ProRS, and Combined scores with clinical models (defined in Supplemental Table S2); compared PRS performance in UKB-PPP with the full UKB cohort; assessed train–test stability of incidence across score deciles; and repeated top-quartile enrichment after excluding events occurring within two years of baseline. Analyses were performed in Python 3.11 using scikit-survival, lifelines, scikit-learn, and SciPy. Code is available at https://github.com/jfieggen/trial_enrichment.

## 3 Results

### 3.1 Study population

After quality control and imputation, 44,802 UKB-PPP participants had measurements for 2,916 plasma proteins. Supplemental Table S3 summarises the ten disease endpoints in the full UKB cohort and the proteomics subset. In the proteomics subset, incident event counts ranged from 116 for Crohn’s disease to 3,088 for atrial fibrillation. Median follow-up was approximately 13.2 years. The study workflow is shown in Fig. 1.

**Figure 1:**
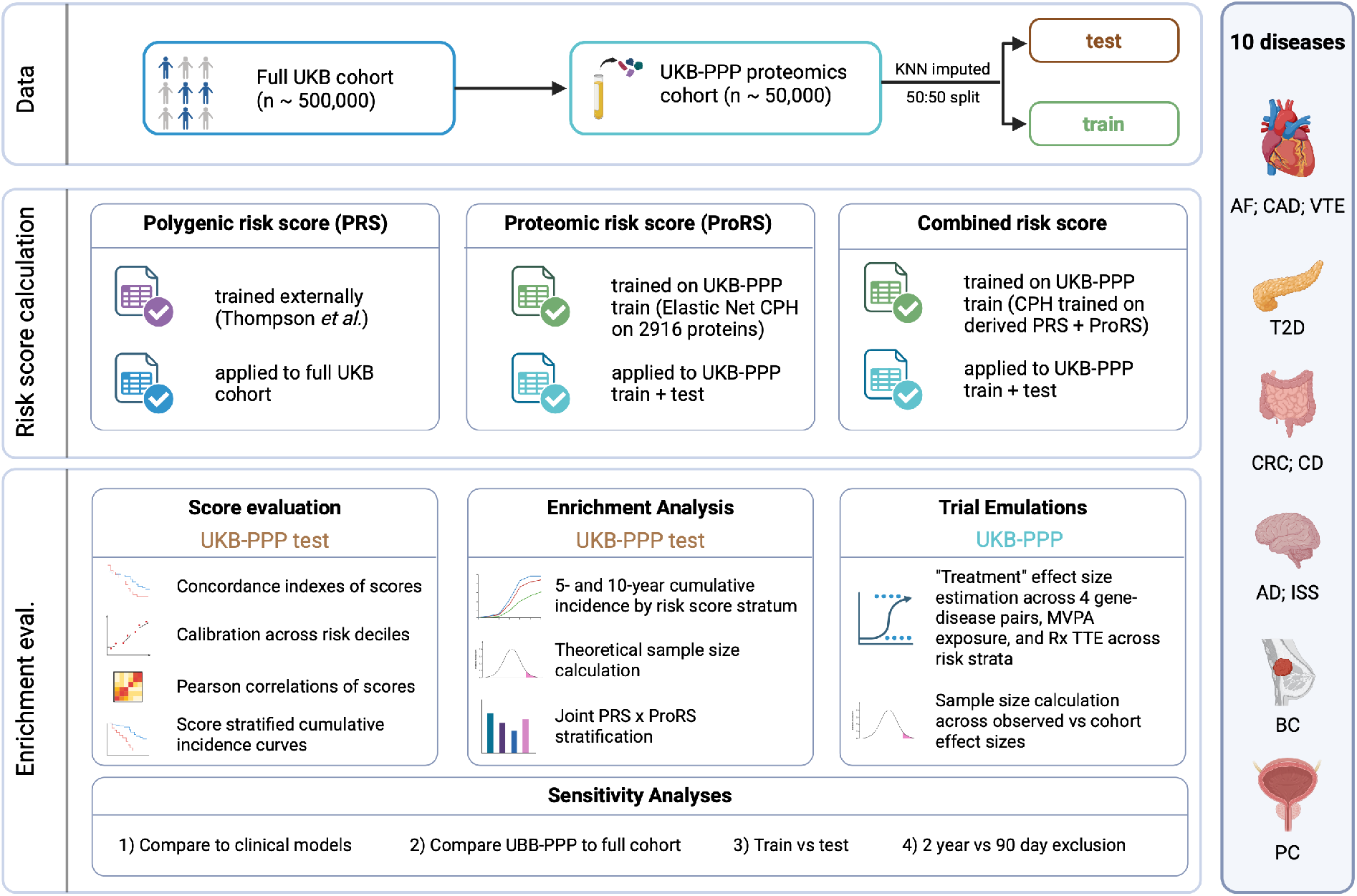
Study design overview. The full UK Biobank (UKB) cohort was subsetted to the UKB UKB-PPP cohort, which underwent k-nearest neighbours (KNN) imputation and a 50:50 split into training and test sets. Proteomic risk scores (ProRS) and Combined risk scores (PRS+ProRS) were trained on the UKB-PPP training set using Cox proportional hazards models and applied to the held-out test data. Risk scores were evaluated for predictive performance, stratification capacity, and theoretical sample size reduction in the test set across 10 incident diseases (right panel). Target trial emulations evaluating genetic, behavioural (MVPA), and pharmacological (Rx) interventions were conducted to assess treatment-like effect stability across enrichment strata. Four sensitivity analyses were undertaken. CAD, coronary artery disease; T2D, type 2 diabetes; AF, atrial fibrillation; BC, breast cancer; CRC, colorectal cancer; PC, prostate cancer; CD, Crohn’s disease; VTE, venous thromboembolism; ISS, ischaemic stroke; AD, Alzheimer’s disease; TTE, target trial emulation; Rx, Pharmaceutical; eval., evaluation; CPH, Cox Proportional Hazards Model; UKB-PPP, UK Biobank pharma proteomics project. Created in https://BioRender.com

### 3.2 Proteomic scores enriched event rates more strongly than PRS

In the held-out test set, ProRS had higher C-indices than PRS for 9 of 10 diseases (Table 1). Median C-index was 0.75 for ProRS and 0.61 for PRS. ProRS performance was highest for Alzheimer’s disease, type 2 diabetes, prostate cancer, atrial fibrillation, ischaemic stroke, and coronary artery disease. Breast cancer was the exception where PRS outperformed ProRS. Combined PRS–ProRS scores gave small additional discrimination over ProRS alone (median ∆C-index = 0.01).

**Table 1:** Discriminative performance and enrichment efficiency of PRS, ProRS, and Combined scores across ten diseases. C-index and potential sample size reduction were evaluated on the held-out test set. Theoretical sample size reductions are reported for the top-quartile stratum (top 25% of the score distribution) at a 10-year endpoint. Calculated sample size reductions assume 20% relative risk reduction and 80% power. 95% confidence intervals are shown in parentheses.

| Disease | C-index |  |  | Sample size reduction (%), Top 25% |  |  |
| --- | --- | --- | --- | --- | --- | --- |
|  | PRS | ProRS | Combined | PRS | ProRS | Combined |
| AD | 0.74 (0.71-0.77) | 0.90 (0.89-0.92) | 0.91 (0.89-0.92) | 63 (58-67) | 74 (73-75) | 74 (73-75) |
| T2D | 0.65 (0.64-0.67) | 0.86 (0.85-0.87) | 0.87 (0.86-0.88) | 41 (36-46) | 71 (70-72) | 71 (70-72) |
| PC | 0.68 (0.66-0.70) | 0.83 (0.81-0.84) | 0.84 (0.83-0.85) | 47 (42-51) | 68 (67-70) | 69 (67-70) |
| AF | 0.61 (0.59-0.62) | 0.80 (0.79-0.81) | 0.81 (0.80-0.82) | 37 (32-41) | 64 (63-66) | 65 (64-67) |
| ISS | 0.57 (0.54-0.60) | 0.75 (0.73-0.78) | 0.76 (0.73-0.78) | 33 (21-43) | 61 (57-64) | 62 (58-65) |
| CAD | 0.58 (0.56-0.59) | 0.74 (0.73-0.75) | 0.76 (0.74-0.77) | 32 (26-37) | 58 (56-60) | 59 (57-61) |
| CD | 0.57 (0.47-0.65) | 0.67 (0.60-0.73) | 0.68 (0.62-0.75) | 31 (-25-54) | 54 (35-64) | 58 (40-67) |
| VTE | 0.60 (0.58-0.62) | 0.68 (0.66-0.70) | 0.71 (0.69-0.73) | 36 (25-44) | 51 (46-55) | 56 (51-60) |
| CRC | 0.61 (0.59-0.64) | 0.65 (0.62-0.67) | 0.69 (0.66-0.71) | 26 (10-35) | 45 (37-52) | 49 (41-54) |
| BC | 0.66 (0.63-0.68) | 0.55 (0.52-0.57) | 0.60 (0.58-0.62) | 45 (37-50) | 22 (11-31) | 32 (22-39) |

AD, Alzheimer’s disease; T2D, type 2 diabetes; PC, prostate cancer; AF, atrial fibrillation; ISS, ischaemic stroke; CAD, coronary artery disease; CD, Crohn’s disease; VTE, venous thromboembolism; CRC, colorectal cancer; BC, breast cancer.

Risk-score enrichment increased observed 10-year cumulative incidence across diseases and score types (Fig. 2; Supplementary Fig. S1). At the top-quartile stratum, ProRS enrichment achieved risk ratios relative to the unselected population of 3.4 for type 2 diabetes, 3.2 for prostate cancer, 2.8 for atrial fibrillation, 2.6 for ischaemic stroke, and 2.4 for coronary artery disease at 10 years. For coronary artery disease, 10-year cumulative incidence rose from 5.1% in the unselected population to 12.1% in the top quartile of the Combined score, and from 3.4% to 11.7% for type 2 diabetes; at the top 5% threshold these reached 21.6% and 30.0% respectively, though at a screening burden of roughly 20 individuals screened per participant enrolled. Under a fixed 20% relative hazard reduction, 10-year endpoint, and 80% power, top-quartile Combined-score enrichment reduced theoretical sample-size requirements by 32–74% across diseases (Table 1). Median top-quartile reduction was 61% for Combined scores, 60% for ProRS, and 37% for PRS. These estimates represent event-driven upper bounds because they do not include screening cost, assay cost, staggered accrual, or possible effect modification.

**Figure 2:**
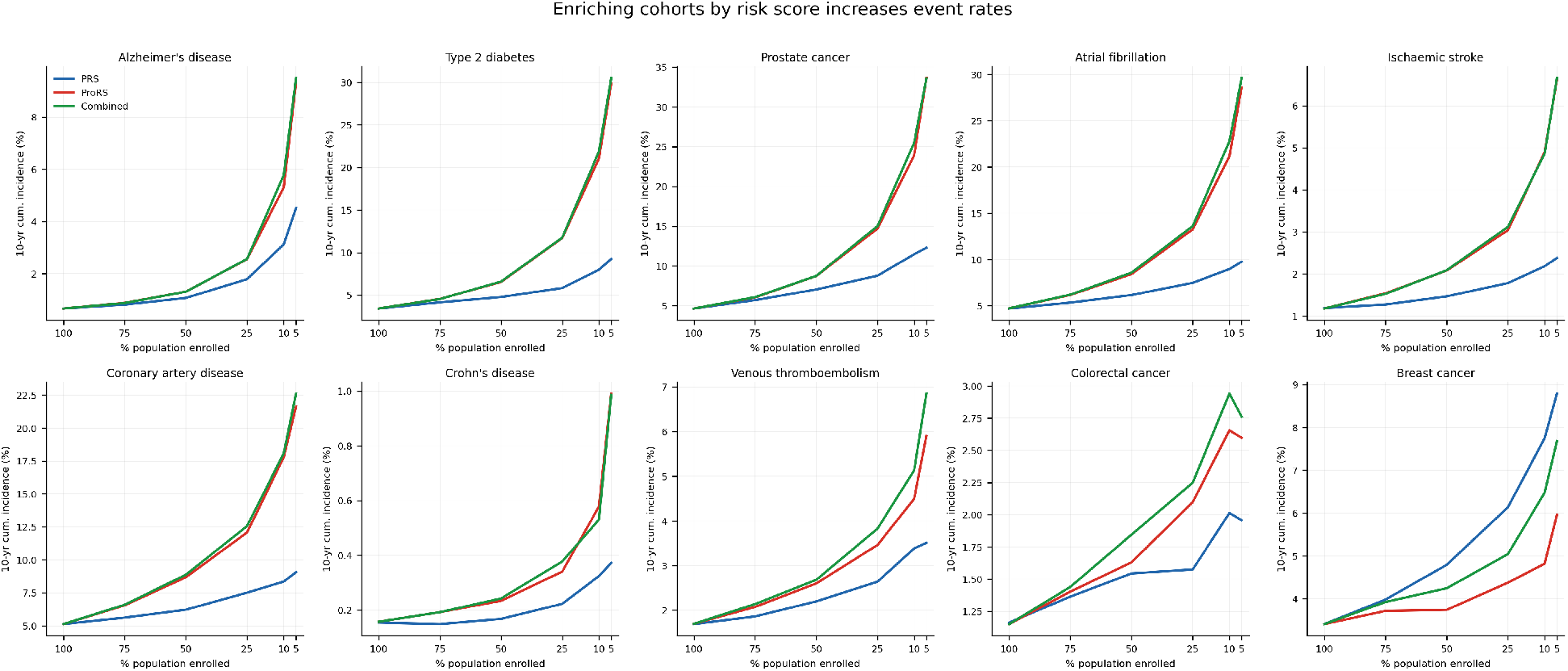
Enriching trial populations by risk score increases event rates. Each panel shows the 10-year cumulative incidence (Kaplan-Meier) at each risk stratum, for PRS (blue), ProRS (red), and Combined (green) scores. Restricting enrolment to higher-risk strata substantially increases the expected event rate.

### 3.3 PRS and ProRS identified partly distinct high-risk populations

PRS and ProRS were weakly correlated across diseases (median Pearson |*r*| = 0.04; Supplementary Fig. S2). The highest correlations were observed for Alzheimer’s disease (*r* = 0.19), prostate cancer (*r* = 0.18), and type 2 diabetes (*r* = 0.17). For the remaining seven diseases, |*r*| *<* 0.07.

Joint top-quartile stratification showed that participants with both high PRS and high ProRS generally had the highest observed incidence (Fig. 3). For type 2 diabetes, 10-year cumulative incidence ranged from 0.6% in the low-PRS/low-ProRS group to 15.6% in the high-PRS/high-ProRS group, with comparable patterns for prostate cancer (1.1% to 21.8%), atrial fibrillation (1.6% to 19.2%), and coronary artery disease (2.5% to 17.0%). For breast cancer, the high-PRS/high-ProRS group reached a higher incidence (7.5%) than the top PRS (6.1%) or Combined (5.4%) quartiles. This pattern suggests that inherited and proteomic risk capture related but non-identical information. Linear Combined scores gave only modest gains over ProRS, suggesting that joint or sequential use of the two modalities may be more informative than a single additive score for pure risk enrichment.

**Figure 3:**
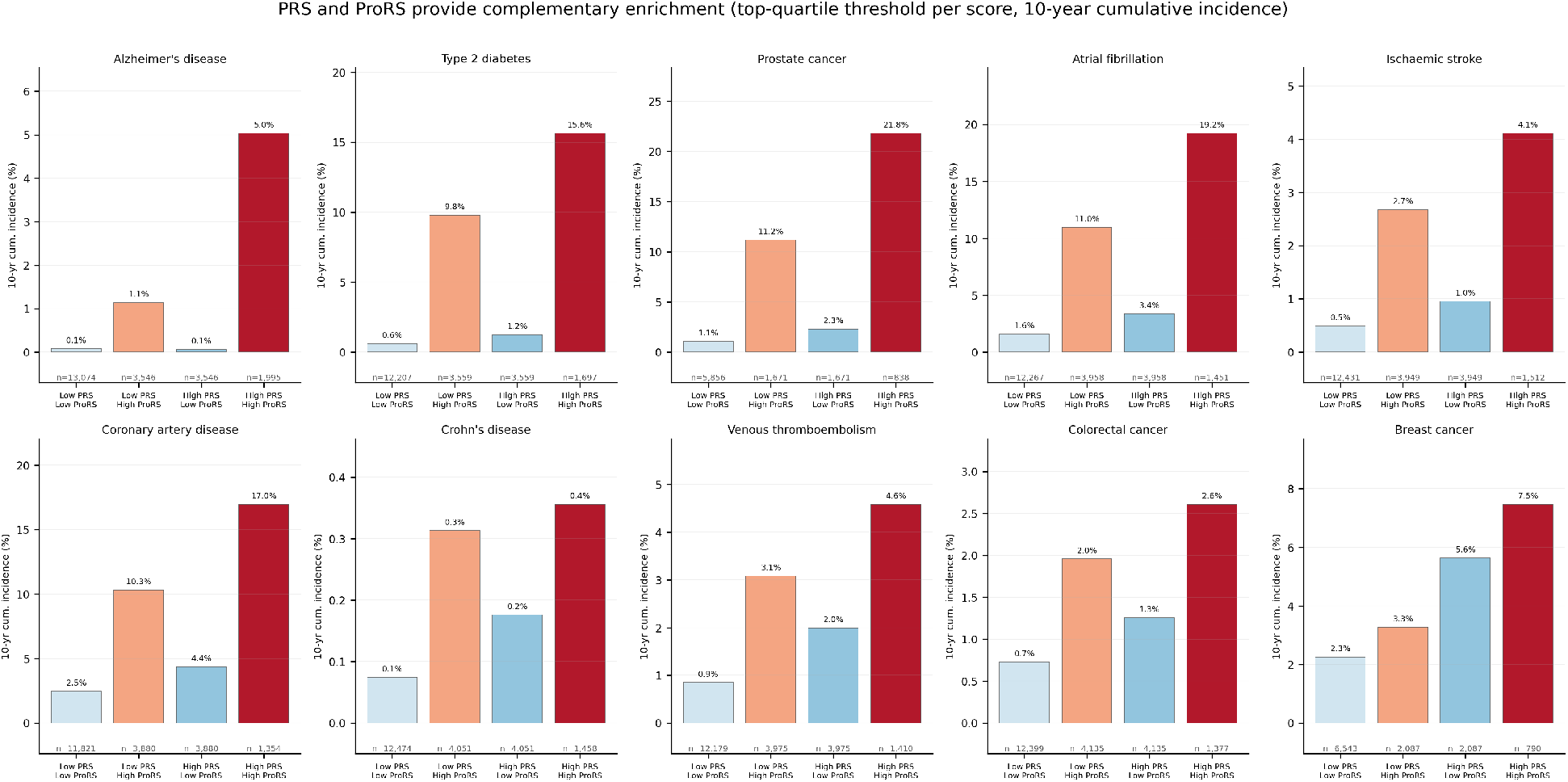
PRS and ProRS provide complementary enrichment. Bar charts show 10-year cumulative incidence within four jointly defined risk groups across ten diseases. The high-PRS/high-ProRS group (dark red) consistently shows the highest incidence.

Stratifying the test data by score decile confirmed increasing observed incidence across deciles, particularly for ProRS and the Combined score. There was more noise in diseases with lower incidence rates and/or lower C-indices (Supplementary Fig. S3).

### 3.4 Event-rate gains did not always imply trial-efficiency gains

We next evaluated whether intervention-analogue effects were preserved across enrichment strata (Fig. 4). Three examples showed broadly preserved protective associations. The *APOE* rs7412 variant remained strongly protective for Alzheimer’s disease at every threshold (aHR range 0.28–0.37, all strata *p <* 0.05), while non-carrier 10-year cumulative incidence rose from 0.8% in the unselected cohort to 2.8% in the top ProRS quartile. The *PCSK9* rs11591147 variant showed a consistently protective direction for coronary artery disease across all strata (aHR range 0.68–0.87), although few individual strata reached significance. High versus low MVPA was also robustly protective for all-cause mortality (aHR range 0.42–0.58 across all score types and strata, all statistically significant).

**Figure 4:**
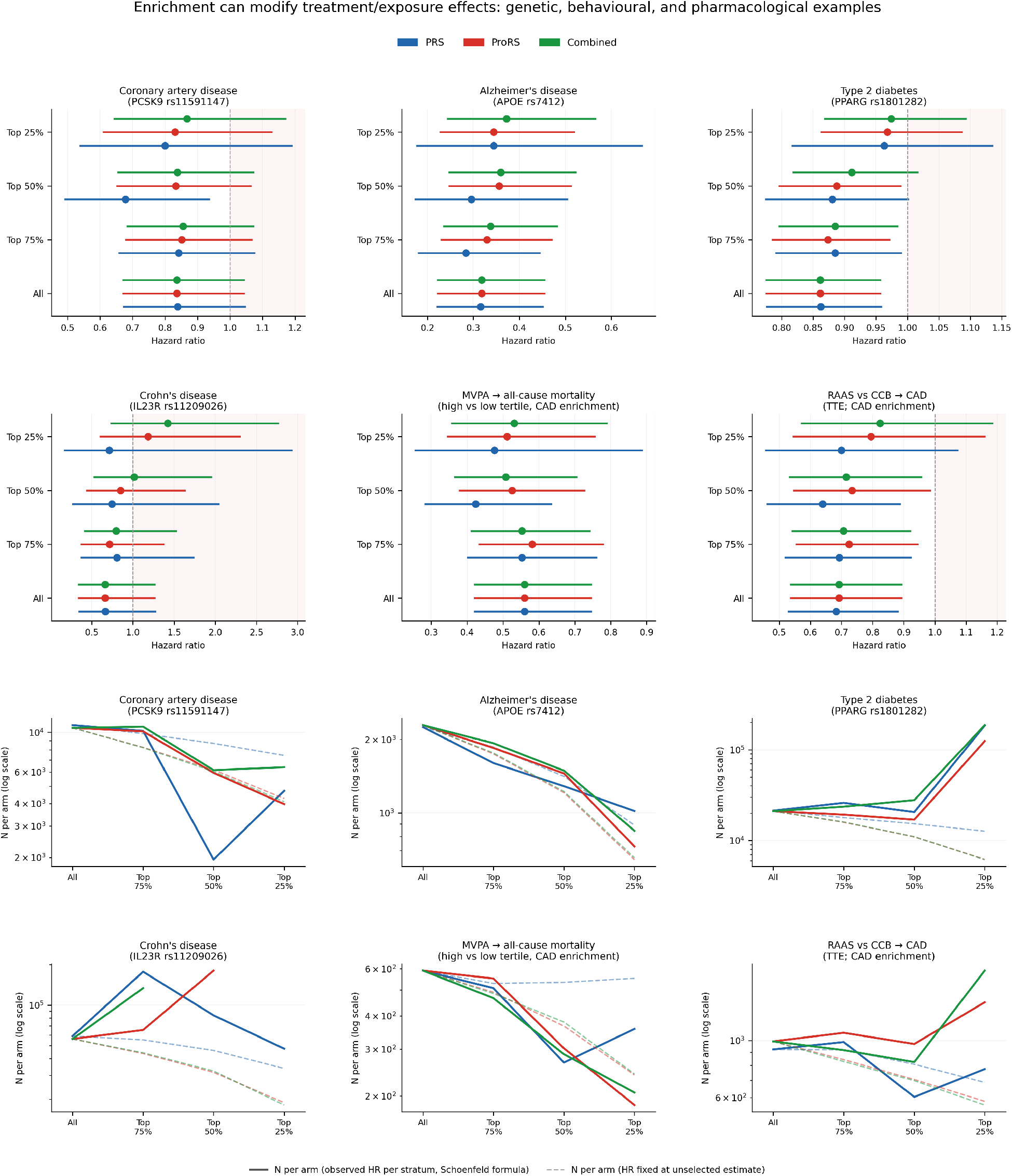
Enrichment can alter intervention-like effects. *Rows 1–2 (forest plots):* Adjusted hazard ratios across enrichment strata for six exposures: four protective genetic variants (carrier vs. non-carrier, adjusted for age and sex; *PCSK9, APOE, PPARG, IL23R*), accelerometer-measured MVPA (high vs. low tertile, adjusted for age, sex, and season of wear), and RAAS vs. CCB initiation (IPTW-weighted Cox, propensity model: age, sex, ethnicity, SBP, eGFR, diabetes, BMI). All scores are restricted to the UKB-PPP data. *Rows 3–4 (N-per-arm):* Required sample size per arm (log scale, 80% power, two-sided *α* = 0.05) as a function of enrichment stratum. Solid lines use the stratum-specific adjusted HR; dashed lines hold the HR fixed at its unselected estimate. Sample size estimates truncated when effect-size reverses rendering a theoretical trial infeasible.

Other examples showed attenuation, with a dependence on score type. The *PPARG* rs1801282 variant, modestly protective for type 2 diabetes in the unselected cohort (aHR = 0.86, 95% CI 0.78–0.96), attenuated to a null effect in the top quartile across all three score types (PRS aHR = 0.96, 95% CI 0.82–1.14; ProRS aHR = 0.97, 95% CI 0.87–1.09; Combined aHR = 0.97, 95% CI 0.87–1.09). For *IL23R* rs11209026 and Crohn’s disease, the effect was score-type-dependent: protective in the unselected cohort (aHR = 0.66, 95% CI 0.34– 1.26), it remained protective under PRS-based enrichment through the top quartile (aHR = 0.71, 95% CI 0.17–2.93) but moved above the null under ProRS (aHR = 1.18, 95% CI 0.61–2.30) and Combined (aHR = 1.42, 95% CI 0.73–2.76) enrichment, although confidence intervals were wide due to the rare endpoint making any firm score-specific assertions impossible. The RAAS-inhibitor versus calcium channel blocker comparison for CAD followed the same pattern: point estimates were attenuated in the most enriched ProRS and Combined strata but broadly preserved under PRS enrichment. These should not be viewed as definitive biological claims for the individual targets but rather indication that a high-risk stratum can have both a higher event-rate and a smaller, or even reversed, intervention-analogue effect, and that this can be related to the type of score used to define it.

These differences changed the hypothetical trial efficacy. When the intervention-analogue effect was preserved, rising event rates reduced theoretically required sample sizes. When the effect attenuated, higher event rates were partly or fully offset by smaller stratum-specific effects. The dashed counterfactual lines in Fig. 4, which assume the unselected-population hazard ratio is constant across strata, show how fixed-effect assumptions can overstate the efficiency of enriched designs.

### 3.5 Sensitivity analyses

Clinical-model comparisons showed that no clinical model exceeded the Combined score for discrimination or theoretical enrichment efficiency, although several disease-specific clinical models approached ProRS or Combined performance and most outperformed PRS (Supplementary Fig. S4). PRS performance was similar in the UKB-PPP test set and the full UKB cohort (Supplementary Table S4). Train–test incidence curves were generally similar across score deciles, with greater instability for lower-incidence endpoints and possible top-decile overfitting for Crohn’s disease and breast cancer ProRS (Supplementary Fig. S5). Excluding events within two years of baseline had little effect on top-quartile incidence or theoretical sample-size reduction (Supplementary Table S5).

## 4 Discussion

We evaluated plasma proteomic risk scores as tools for prognostic enrichment in prevention-oriented trials. ProRS identified substantially higher-risk populations than PRS for most diseases and produced large theoretical sample-size reductions under fixed-effect assumptions. However, the intervention-analogue analyses showed that event-rate enrichment and trial-efficiency enrichment can diverge. In some examples, the estimated effect was preserved across enriched strata; in others, it attenuated, reducing or eliminating the expected efficiency gain.

### 4.1 Proteomic biomarkers as trial-enrichment tools

The results support the use of plasma proteins as scalable enrichment variables. ProRS outperformed PRS for 9 of 10 endpoints and concentrated events in high-score strata. This is consistent with the biological basis of the circulating proteome – unlike PRS, it can reflect inherited predisposition, acquired risk factors, inflammation, metabolic status, and subclinical disease processes. The exception of breast cancer, where PRS performed better than ProRS, is consistent with its strong germline contribution to risk [28] and highlights that the relative value of each modality is disease-dependent. The low correlation between PRS and ProRS could also enable a serial screening strategy, in which low-cost genotyping is performed first and proteomic assays are reserved for the PRS-enriched subset, potentially reducing screening costs.

The clinical-model comparison is important to note. Although Combined scores performed best overall, several disease-specific clinical models approached ProRS performance. However, one can anticipate clinical risk models, especially those that contain blood-based biomarkers, to have similar limitations to proteomic risk scores.

### 4.2 Relation to PRS-guided enrichment frameworks

Our study builds on PRS-guided in silico enrichment work. Previous work has used protective genetic variants as therapeutic analogues and showed that PRS restriction can increase disease prevalence, improve power, and reduce theoretical sample size [18]. We extend this in two ways. First, we evaluate proteomic rather than only inherited risk, and compare PRS, ProRS, and their combination across ten diseases. Second, we separate event-rate enrichment from effect preservation. This distinction is particularly important for phenotypic biomarkers because they may identify people at a different biological stage of disease rather than only people with higher lifelong susceptibility. *PPARG* /T2D saw a similar attenuation pattern in PRS and ProRS, while other examples (e.g. *IL23R*/CD) saw potentially discordant alterations in observed enriched-stratum effect size underscoring this point.

The low correlation between PRS and ProRS indicated that these two scores often identified different individuals, and the highest-risk joint strata generally had the highest observed incidence. This complementarity may be useful for trial design, but it also means that different enrichment strategies will define different target populations. This is crucial when considering the anticipated mechanism of the intervention and thus ensuring a trial is not prognostically enriched for individuals less susceptible to that mechanism.

### 4.3 Mechanism-aware enrichment

The main practical implication is that prognostic enrichment should be mechanism-aware. Standard sample-size calculations assume that the relative effect is stable across risk strata. That assumption was plausible in some of our examples: *APOE* /AD and MVPA/mortality showed preserved effects, and *PCSK9* /CAD remained directionally protective. In these settings, theoretical prognostic enrichment worked as expected, with higher event rates translating into smaller theoretical trials.

Other examples showed why the assumption should be considered carefully before prognostic enrichment is considered. *PPARG* /T2D attenuated toward the null in enriched strata, and *IL23R*/Crohn’s disease was compatible with attenuation under ProRS-based enrichment, although estimates were imprecise. These findings are not a comment on the biological claims for those targets but rather illustrative that a high-risk stratum may accidentally select for individuals for whom a theoretical preventative therapy is less beneficial. This problem may be particularly relevant for proteomic enrichment. A proteomic score may be excellent for predicting events but less appropriate for selecting participants for an intervention that acts before those downstream changes occur. This is important given the ongoing discourse around protein-based biomarkers potentially enabling prevention in diseases such as Alzheimer’s [29].

### 4.4 Implications for prevention-trial design

Our findings suggest a staged approach to biomarker-guided enrichment. First, investigators should quantify the event-rate gain achieved by the candidate enrichment variable. Second, they should evaluate whether the intervention mechanism is expected to operate in the enriched population, using prior biology, human genetics, completed trials, and/or target-trial emulation in biobanks [30] where possible. Third, they should account for screening burden and assay cost. Finally, unless effect preservation is well supported, biomarker-defined groups may be safer as pre-specified strata, adaptive-enrichment variables, or health-economic subgroups rather than as restrictive eligibility criteria.

Additional concerns with prognostic enrichment remain unaddressed. Excluding lower-risk individuals can reduce generalisability and prognostic enrichment does not reduce the number needed to screen, so recruitment burden – often the key constraint in large trials – is largely unchanged [31]. This further underscores the overarching importance of inclusive trial designs as the default with enrichment only being considered in carefully selected cases where broad trials may not be pragmatic.

### 4.5 Limitations

This study has important limitations. UK Biobank is affected by healthy-volunteer selection and is predominantly of European ancestry, limiting generalisability of the findings in this study [32]. ProRS were developed and evaluated within the same cohort and external validation should be considered (although previous literature has demonstrated that proteomic risk scores generalise well [9] providing some reassurance). The sample-size calculations are theoretical and exclude screening costs, assay costs, staggered recruitment, adherence, competing risks, and operational constraints. Genetic variants, MVPA, and antihypertensive comparisons are imperfect analogues of randomised interventions. Relatedly, estimating an exposure–outcome association within a score-defined stratum conditions on a variable that is plausibly a common effect of the exposure and the outcome’s other determinants (especially for the genetic analogues) so this conditioning can open a collider path and distort the carrier–outcome estimate independent of any true effect modification. The intervention-analogue analyses were limited by the size of the proteomics subset, and estimates for rare endpoints such as Crohn’s disease were imprecise. Finally, plasma protein concentrations are dynamic, so timing of measurement may affect enrichment performance.

### 4.6 Conclusions

Proteomic risk scores can identify high-event-rate populations and may substantially reduce theoretical sample-size requirements for large-scale prevention-oriented trials. However, prognostic enrichment faces many important challenges – a key one of which is considering whether the relevant intervention effect will be preserved in the enriched group. Proteomic and polygenic scores may therefore be most useful as mechanism-aware stratification or adaptive-design variables, rather than as default restrictive eligibility criteria.

## Supporting information

tripodAI checklist

## Data Availability

This research was conducted using the UK Biobank Resource (application number 83801). UK Biobank data are available to approved researchers via the UK Biobank (https://www.ukbiobank.ac.uk). Analysis code is available at https://github.com/jfieggen/trial_enrichment.

## Author Contributions

J.F. and L.C. conceptualized the study. J.F. performed the data analysis and wrote the first draft of the manuscript. L.C., D.A.C., and C.C.B. supervised the research. All authors (J.F., G.S., A.N., B.M.S., A.T., C.C.B., D.A.C., L.C.) contributed to the interpretation of the results, critically reviewed and revised the manuscript, and approved the final version for submission.

## Acknowledgements

This research has been conducted using the UK Biobank Resource under Application Number 83801. This work uses data provided by patients and collected by the NHS as part of their care and support. We thank the participants of the UK Biobank study without whom this research would not have been possible. Computing for this study used the Oxford Biomedical Research Computing (BMRC) facility, a joint development between the Wellcome Centre for Human Genetics and the Big Data Institute supported by Health Data Research UK and the NIHR Oxford Biomedical Research Centre. JF and BS are supported by Rhodes Scholarships. DAC is funded by an National Institute for Health Research (NIHR) Research Professorship; a Royal Academy of Engineering Research Chair; and InnoHK Hong Kong Centre for Cerebro-cardiovascular Engineering (COCHE) and supported by the Pandemic Sciences Institute at the University of Oxford; the NIHR Oxford Biomedical Research Centre (BRC); and the EPSRC (grant EP/W031744/1). LC is supported by the National Institute for Health Research (NIHR) Oxford Biomedical Research Centre (BRC); NIHR Applied Research Collaboration (ARC) Oxford and Thames Valley at Oxford Health NHS Foundation Trust; and the Nuffield Department of Primary Care Health Sciences at the University of Oxford. The views expressed are those of the authors and not necessarily those of the NHS, the NIHR or the Department of Health and Social Care.

