## supplemental figs and tables 1-5 for "Beyond event-rate enrichment: proteomic risk scores for mechanism-aware prevention trial design"

### Supplementary Information

#### Supplementary Methods S1: Theoretical sample-size calculation

For each disease, score type, and enrichment stratum we estimated the number of participants a hypothetical two-arm, parallel-group, time-to-event trial would require to detect a fixed relative treatment effect. As enrichment raises the event rate, fewer participants are required to accrue the events needed for a given power. These calculations are theoretical upper bounds: they assume instantaneous enrolment, no drop-out beyond what is captured by the Kaplan–Meier estimator, no screening or operational cost, and – unless stated otherwise – a relative treatment effect that is constant across risk strata, an assumption we revisit in the effect-preservation analyses.

Required sample sizes were approximated using an event-driven log-rank design under the proportional-hazards assumption. The total number of events required across both arms was obtained from the Schoenfeld formula [1]:

$$D = \frac{(z_{\alpha/2} + z_\beta)^2}{P_A P_B \, (\log \theta)^2}, \tag{1}$$

where $D$ is the total required number of events, $\theta$ is the target hazard ratio, $P_A$ and $P_B$ are the allocation proportions ($P_A + P_B = 1$), $z_{\alpha/2} = 1.96$ for two-sided $\alpha = 0.05$, and $z_\beta$ corresponds to the desired power. We assumed 1:1 randomisation ($P_A = P_B = 0.5$) and specified $\theta = 0.80$ (a 20% relative hazard reduction) as the primary scenario.

For each stratum, the control-arm cumulative incidence at the target horizon, $p_c$, was set to the observed Kaplan–Meier estimate within that stratum, so censoring and loss to follow-up enter through the KM estimator. Under proportional hazards the treatment-arm incidence is

$$p_t = 1 - (1 - p_c)^\theta. \tag{2}$$

With $N$ participants per arm, the expected number of events across both arms by the horizon is $N(p_c + p_t)$. Equating this to $D$ gives

$$n_{\text{per arm}} = \left\lceil \frac{D}{p_c + p_t} \right\rceil. \tag{3}$$

Calculations assumed instantaneous enrolment, so each randomised participant contributes the full follow-up duration to the target horizon. Sample-size reductions were expressed as the percentage change in $n_{\text{per arm}}$ relative to the unselected population at the same horizon and power, with 80% power and a 10-year endpoint as the primary comparison. Full results across horizons (5- and 10-year), relative risk reductions (15%, 20%, 25%), and power levels (80%, 90%) are given in Supplementary Table S7. Ninety-five percent confidence intervals for both absolute $n_{\text{per arm}}$ and the percentage reduction were obtained from 300 bootstrap resamples.

#### Supplementary Figure S1: Kaplan-Meier curves by risk quartiles

Kaplan-Meier curves with 95% confidence intervals across risk quartiles for PRS, ProRS, and Combined scores for all 10 evaluated diseases.

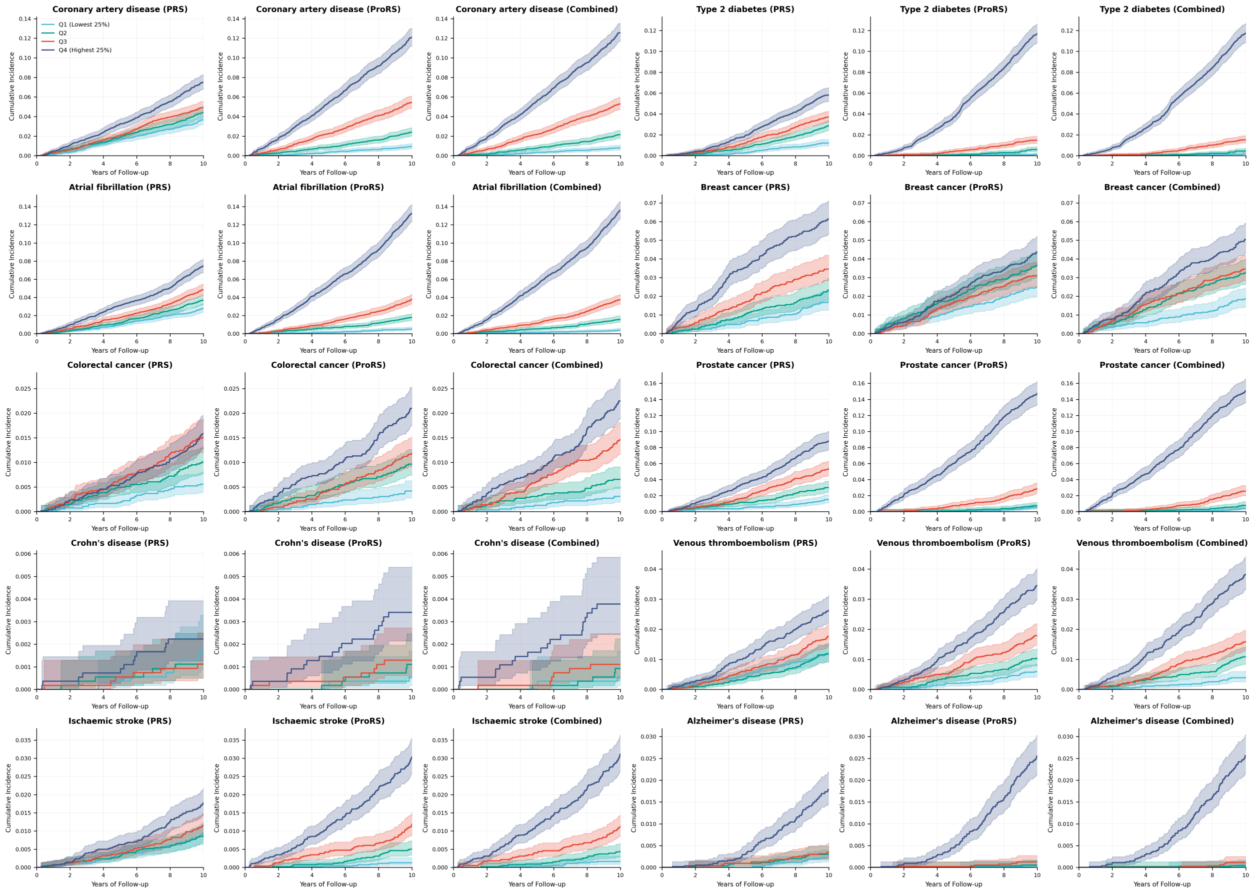


Figure S1

### Supplementary Figure S2: PRS–ProRS correlation

Pearson correlation coefficients between PRS and ProRS for each of the ten diseases. Most correlations are near zero, confirming that the two scores capture largely independent risk dimensions. The highest correlations are observed for Alzheimer's disease ($r = 0.19$), prostate cancer ($r = 0.18$), and type 2 diabetes ($r = 0.17$).

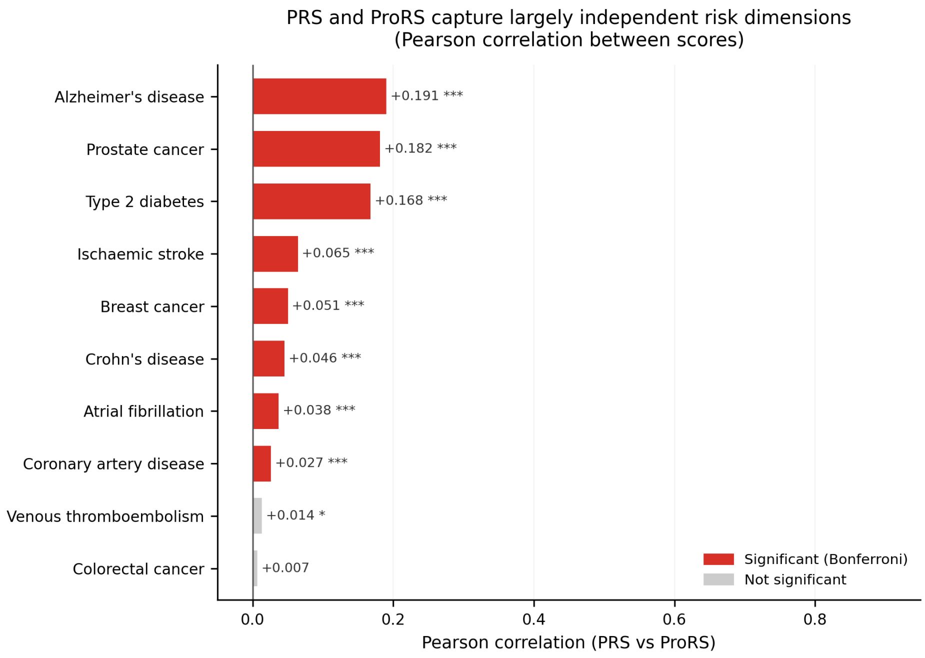


Figure S2

#### Supplementary Figure S3: Incidence by risk decile

Observed 10-year cumulative incidence versus mean predicted risk score within deciles of PRS, ProRS, and Combined scores, for each disease. ProRS and Combined scores show consistent risk increases for most diseases.

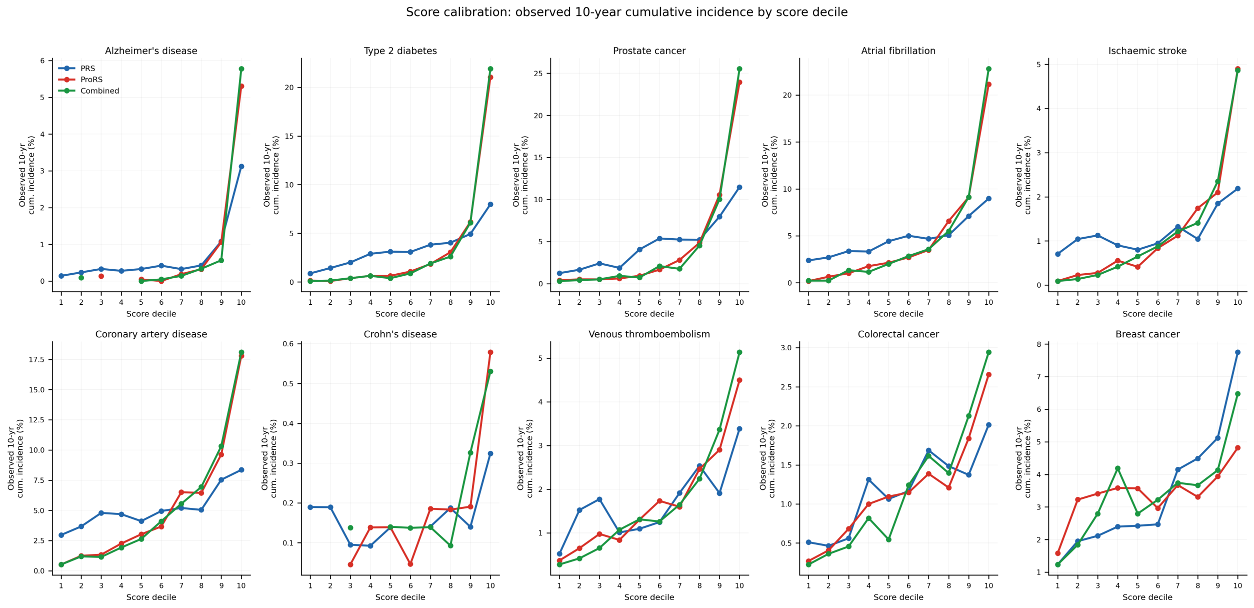


Figure S3

#### Supplementary Figure S4: Comparison of clinical and -omics based enrichment scores in terms of discrimination and theoretical sample size reduction.

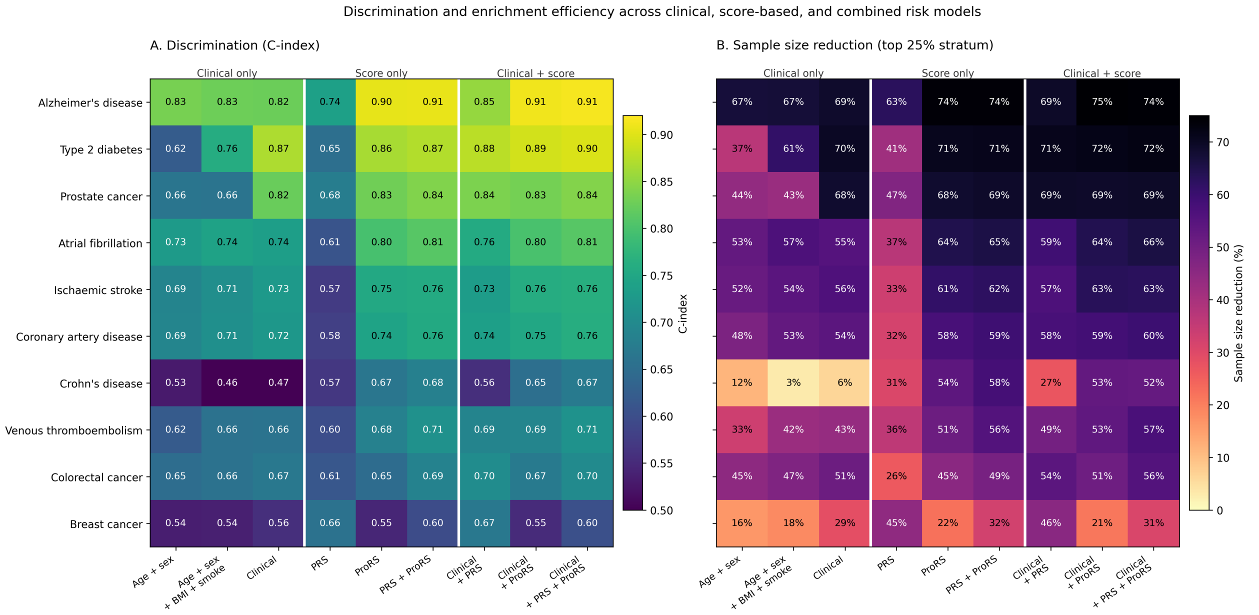


Figure S4

#### Supplementary Figure S5: Calibration of train and test score-based enrichment

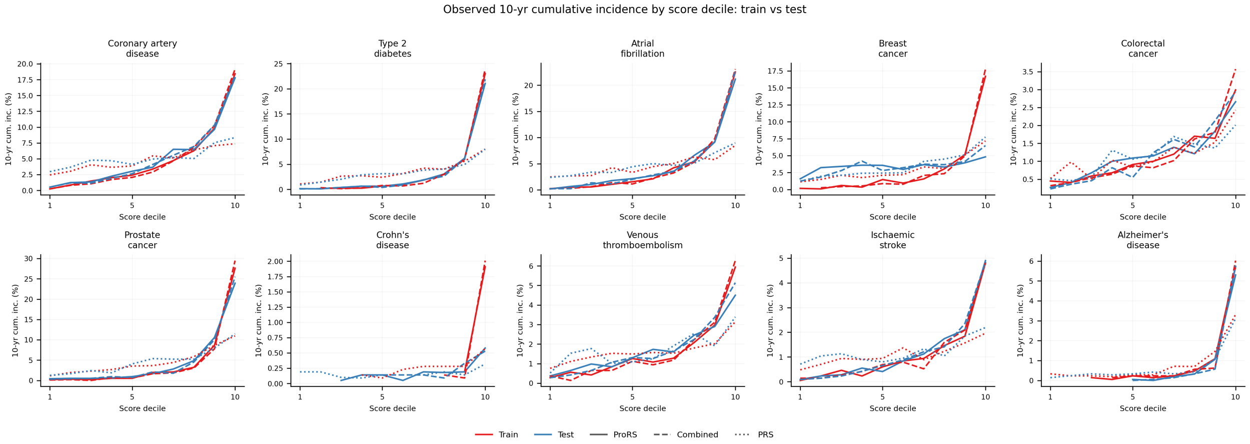


Figure S5

### Supplementary Table S1: Disease definitions

Table S1: ICD-10, ICD-9, data sources, sex restrictions, and self-reported disease codes used for defining incident disease endpoints and prevalent-case exclusions.

| Disease Endpoint | ICD-10 Codes | ICD-9 Codes | Data Sources | Sex | Self-Report Codes[a] |
|---|---|---|---|---|---|
| Coronary artery disease (CAD) | I21, I22, I23, I25 (excl. I25.2) | 410–414 | Hospital | None | 1074, 1075 |
| Type 2 diabetes (T2D) | E11 | 250.0, 250.2, 250.4, 250.6, 250.8 | Hospital | None | 1220, 1223 |
| Atrial fibrillation (AF) | I48 | 427.3 | Hospital | None | 1077, 1471 |
| Breast cancer (BC) | C50 | 174 | Hospital, Cancer | Female | 1002[b] |
| Colorectal cancer (CRC) | C18, C19, C20 | 153, 154 | Hospital, Cancer | None | 1020, 1021, 1022, 1023[b] |
| Prostate cancer (PC) | C61 | 185 | Hospital, Cancer | Male | 1044[b] |
| Crohn's disease (CD) | K50 | 555 | Hospital | None | 1459, 1461, 1462 |
| Venous thromboembolism (VTE) | I26, I80.1, I80.2, I80.3, I82 | 415–417, 451, 453 | Hospital | None | 1068, 1093, 1094 |
| Ischaemic stroke (ISS) | I63 | 434, 436 | Hospital | None | 1081, 1583 |
| Alzheimer's disease (AD) | G30, F00 | 331.0 | Hospital | None | 1263 |

[a] UK Biobank non-cancer self-reported disease codes (Field 20002) used for prevalent-case exclusion at baseline.

[b] UK Biobank cancer self-reported disease codes (Field 20001) used for prevalent-case exclusion at baseline.

Hospital = Hospital Inpatient Data; Cancer = National Cancer Register; ICD = International Classification of Diseases

#### Supplementary Table S2: Disease-Specific Clinical Model Variables.

Table S2: Overview of Disease-Specific Clinical Model Variables and UK Biobank (UKB) Field IDs

| Disease | Included Clinical Covariates (UKB Field ID) |
|---|---|
| **CAD** | Age, Sex, BMI, Smoking, Systolic BP (4080), LDL (30780), Prevalent T2D, FH Heart Disease (20107, 20110, 20111; code 1) |
| **T2D** | Age, Sex, BMI, Smoking, HbA1c (30750), FH Diabetes (20107, 20110, 20111; code 9) |
| **AF** | Age, Sex, BMI, Systolic BP (4080), Alcohol Frequency (1558) |
| **ISS** | Age, Sex, Smoking, Systolic BP (4080), Prevalent T2D, Prevalent AF, FH Stroke (20107, 20110, 20111; code 2) |
| **VTE** | Age, Sex, BMI, Smoking, Hormone use - Oral Contraceptives (2784) or HRT (2814) |
| **BC** | Age, BMI, FH Breast Cancer (20107, 20110, 20111; code 5) |
| **PC** | Age, FH Prostate Cancer (20107, 20110, 20111; code 13), KLK3 |
| **CRC** | Age, Sex, BMI, Smoking, Alcohol Frequency (1558), FH Bowel Cancer (20107, 20110, 20111; code 4) |
| **CD** | Age, Sex, Smoking |
| **AD** | Age, Sex, Townsend Deprivation Index (22189), FH Alzheimer's/Dementia (20107, 20110, 20111; code 10) |

Universal covariates included age at recruitment (21022), sex (31), BMI (21001), and smoking status (previous/current; 20116). FH, Family History. UKB Field IDs 20107, 20110, and 20111 correspond to the illnesses of the father, mother, and siblings, respectively. KLK3 was used as a PSA proxy from the proteomics data. These variables were used for the "Clinical" models described in Fig. S4.

#### Supplementary Table S3: Summary of disease endpoints defined in the full UK Biobank cohort and the proteomics subset.

Table S3: N eligible refers to participants remaining after prevalent-case exclusion. Incidence is the proportion of eligible participants with an incident event during follow-up.

| | | Full UKB Cohort | | | | Proteomics Subset | | |
|---|---|---|---|---|---|---|---|---|
| **Disease** | **Sex** | **N eligible** | **N prevalent** | **N incident** | **Inc. (%)** | **N eligible** | **N incident** | **Inc. (%)** |
| CAD | All | 476,706 | 25,223 | 31,936 | 6.70 | 42,332 | 2,945 | 6.96 |
| T2D | All | 477,629 | 24,300 | 23,757 | 4.97 | 42,470 | 2,208 | 5.20 |
| AF | All | 492,329 | 9,600 | 33,447 | 6.79 | 43,756 | 3,088 | 7.06 |
| BC | Female | 262,752 | 10,279 | 11,700 | 4.45 | 23,349 | 1,002 | 4.29 |
| CRC | All | 499,159 | 2,770 | 8,401 | 1.68 | 44,552 | 747 | 1.68 |
| PC | Male | 225,292 | 3,606 | 14,424 | 6.40 | 20,243 | 1,299 | 6.42 |
| CD | All | 498,703 | 3,226 | 1,292 | 0.26 | 44,488 | 116 | 0.26 |
| VTE | All | 488,704 | 13,225 | 12,089 | 2.47 | 43,518 | 1,136 | 2.61 |
| ISS | All | 495,314 | 6,615 | 8,628 | 1.74 | 44,141 | 828 | 1.88 |
| AD | All | 501,808 | 121 | 4,086 | 0.81 | 44,774 | 568 | 1.27 |

CAD, coronary artery disease; T2D, type 2 diabetes; AF, atrial fibrillation; BC, breast cancer; CRC, colorectal cancer; PC, prostate cancer; CD, Crohn's disease; VTE, venous thromboembolism; ISS, ischaemic stroke; AD, Alzheimer's disease. Inc., incidence.

#### Supplementary Table S4: PRS performance in the UKB-PPP proteomics subset compared with the full UK Biobank cohort.

Table S4: Comparison of polygenic risk score (PRS) discrimination and theoretical enrichment efficiency in the held-out UKB-PPP test set ($n \approx 22{,}000$) and the full UK Biobank cohort ($n \approx 500{,}000$). C-index and sample-size-reduction estimates are similar across the two populations suggesting that the proteomics subset is broadly representative of the full cohort for the purposes of PRS-based enrichment evaluation.

| | **UKB-PPP test set** | | | | **Full UKB cohort** | | | |
|---|---|---|---|---|---|---|---|---|
| **Disease** | **N** | **Events** | **C-index** | **SS red. (%)** | **N** | **Events** | **C-index** | **SS red. (%)** |
| AD | 22,161 | 273 | 0.74 | 63 | 485,596 | 3,910 | 0.70 | 57 |
| T2D | 21,022 | 1,054 | 0.65 | 41 | 462,379 | 22,780 | 0.65 | 44 |
| PC | 10,036 | 638 | 0.68 | 47 | 218,837 | 14,052 | 0.69 | 50 |
| AF | 21,634 | 1,543 | 0.61 | 37 | 476,404 | 32,329 | 0.62 | 39 |
| ISS | 21,841 | 430 | 0.57 | 33 | 479,360 | 8,340 | 0.57 | 26 |
| CAD | 20,935 | 1,455 | 0.58 | 32 | 461,373 | 30,844 | 0.59 | 32 |
| CD | 22,034 | 57 | 0.57 | 31 | 482,592 | 1,242 | 0.63 | 41 |
| VTE | 21,539 | 562 | 0.60 | 36 | 472,997 | 11,613 | 0.61 | 37 |
| CRC | 22,046 | 375 | 0.61 | 26 | 483,037 | 8,123 | 0.61 | 37 |
| BC | 11,507 | 528 | 0.66 | 45 | 253,794 | 11,304 | 0.65 | 44 |

SS red., theoretical sample-size reduction at the top-quartile enrichment stratum, computed under a 10-year endpoint, 20% relative hazard reduction, two-sided $\alpha = 0.05$, and 80% power. Disease abbreviations: AD, Alzheimer's disease; T2D, type 2 diabetes; PC, prostate cancer; AF, atrial fibrillation; ISS, ischaemic stroke; CAD, coronary artery disease; CD, Crohn's disease; VTE, venous thromboembolism; CRC, colorectal cancer; BC, breast cancer.

#### Supplementary Table S5: Sensitivity analysis excluding incident events occurring within two years of baseline.

Table S5: Sensitivity analysis re-computing 10-year cumulative incidence and theoretical sample-size reductions after excluding any incident events occurring within two years of baseline (as compared to 90 days). Sample-size reductions are robust to early-event exclusion across all three score types and ten diseases suggesting that proteomic-based enrichment is not driven by occult prevalent disease at baseline.

| | | | Top-25% 10-yr cum. inc. (%) | | SS reduction (%) | |
|---|---|---|---|---|---|---|
| **Disease** | **Score** | **Events <2 yr** | **Original** | **2-yr excl.** | **Original** | **2-yr excl.** |
| AD | PRS | 6 | 1.79 | 1.74 | 63 | 63 |
| AD | ProRS | 6 | 2.56 | 2.48 | 74 | 74 |
| AD | Combined | 6 | 2.56 | 2.47 | 74 | 74 |
| T2D | PRS | 47 | 5.83 | 5.43 | 41 | 41 |
| T2D | ProRS | 47 | 11.71 | 10.99 | 71 | 71 |
| T2D | Combined | 47 | 11.75 | 11.04 | 71 | 71 |
| PC | PRS | 56 | 8.78 | 7.87 | 47 | 48 |
| PC | ProRS | 56 | 14.70 | 12.84 | 68 | 68 |
| PC | Combined | 56 | 15.02 | 13.08 | 69 | 69 |
| AF | PRS | 120 | 7.48 | 6.54 | 37 | 36 |
| AF | ProRS | 120 | 13.26 | 11.80 | 64 | 65 |
| AF | Combined | 120 | 13.62 | 12.21 | 65 | 66 |
| ISS | PRS | 28 | 1.78 | 1.64 | 33 | 35 |
| ISS | ProRS | 28 | 3.04 | 2.70 | 61 | 61 |
| ISS | Combined | 28 | 3.12 | 2.81 | 62 | 63 |
| CAD | PRS | 146 | 7.51 | 6.47 | 32 | 31 |
| CAD | ProRS | 146 | 12.08 | 10.42 | 58 | 57 |
| CAD | Combined | 146 | 12.56 | 10.83 | 59 | 59 |
| CD | PRS | 4 | 0.22 | 0.19 | 31 | 27 |
| CD | ProRS | 4 | 0.34 | 0.30 | 54 | 54 |
| CD | Combined | 4 | 0.38 | 0.32 | 58 | 57 |
| VTE | PRS | 34 | 2.64 | 2.38 | 36 | 36 |
| VTE | ProRS | 34 | 3.46 | 3.17 | 51 | 51 |
| VTE | Combined | 34 | 3.83 | 3.44 | 56 | 55 |
| CRC | PRS | 38 | 1.58 | 1.34 | 26 | 26 |
| CRC | ProRS | 38 | 2.10 | 1.74 | 45 | 44 |
| CRC | Combined | 38 | 2.25 | 1.89 | 49 | 48 |
| BC | PRS | 68 | 6.14 | 4.91 | 45 | 42 |
| BC | ProRS | 68 | 4.38 | 3.78 | 22 | 25 |
| BC | Combined | 68 | 5.04 | 4.28 | 32 | 34 |

Events <2 yr is a count of incident events that occurred within two years of baseline assessment and were excluded in the sensitivity analysis. Theoretical sample size (SS) reduction is estimated as previously described.

#### Large tables: see excel supplement

#### Supplementary Table S6: Full incidence by threshold

Complete table of enrichment metrics (cumulative incidence, incidence rate, eligible fraction, screening burden) across all score types, diseases, and enrichment thresholds (All, Top 75%, 50%, 25%, 10%, 5%). This includes source data for Fig. 2.

#### Supplementary Table S7: Full sample size by threshold

Required sample sizes per arm for all combinations of disease, score type, enrichment stratum, endpoint horizon (5-year, 10-year), relative risk reduction (15%, 20%, 25%), and power (80%, 90%). This includes the source data for Table 1.

#### Supplementary Table S8: Carrier Cox results by stratum

Hazard ratios (with 95% confidence intervals and $p$-values) for carrier status within each enrichment stratum, for all four gene-disease pairs and three score types. This includes the source data for the top two rows of Fig. 4.

#### Supplementary Table S9: Carrier arm absolute risks

10-year cumulative incidence, incidence rate, and person-years for carrier and non-carrier arms within each enrichment stratum. This is the source data for the bottom two rows of Fig. 4.

#### Supplementary Table S10: Joint PRS–ProRS strata

Sample sizes, event counts, and 10-year cumulative incidence within each of the four jointly defined risk groups (low/low, low/high, high/low, high/high) for all ten diseases. This is the source data for Fig. 3.

#### Supplementary Table S11: Calibration by decile

Observed 10-year cumulative incidence and mean score value within each score decile for all diseases and score types. This includes the source data for Fig. S3.

#### Supplementary Table S12: ProRS Coefficients

ProRS disease-specific coefficients estimated in the training data by the Elastic-Net Cox model.

#### Supplementary Table S13: Comparison of clinical and -omics based enrichment scores

Comparison of performance in terms of discrimination and theoretical sample size reduction. This includes the source data for Fig. S4.
